# Predictors of maternal mental health and coping during the COVID-19 pandemic: A multi-country cross-sectional study

**DOI:** 10.64898/2026.05.25.26353920

**Authors:** Chuwen Liu, Mengyun Liu, Sarah Dib, Milagros Ferrando, Masaharu Kagawa, Krongporn Ongprasert, Emeline Rougeaux, Nurul Husna M Shukri, Adriana Vazquez, Jonathan Wells, Mary Fewtrell, Jinyue Yu

**Affiliations:** Childhood Nutrition Research Group, Population, Policy & Practice Department, UCL Great Ormond Street Institute of Child Health, London, UK; Leicester Diabetes Centre and Leicester NIHR Biomedical Research Centre, University of Leicester, Leicester General Hospital, Gwendoline Road, Leicester LE5 4PW, UK; MG. Milagros Ferrando, Servicio de Nutrición, Hospital Mira y López, Santa Fe, Argentina. Facultad de Bioquímica, Universidad Nacional del Litoral, Santa Fe, Argentina; Institute of Nutrition Sciences, Kagawa Nutrition University, Saitama, Japan; Department of Community Medicine, Chiang Mai University, Thailand; Department of Nutrition, Faculty of Medicine & Health Sciences, Universiti Putra Malaysia, Malaysia; Evidence Synthesis Group, Bristol Medical School, University of Bristol, Bristol, UK

**Author notes:** Corresponding: Jinyue Yu, Childhood Nutrition Research Group, Population, Policy & Practice Department, UCL Great Ormond Street Institute of Child Health, London, UK. These two authors contribute equally to this article (Chuwen Liu, Mengyun Liu).

**Keywords:** Mental Health, COVID-19, Postpartum Period, Coping Behaviour, Social Support, Cross-Cultural Comparison

## Abstract

**Objectives and study:** This study aimed to examine predictors of post-partum maternal mental health (MMH) and coping during COVID-19 lockdown across seven countries (the UK, China, Japan, Malaysia, Mexico, Argentina, and Thailand).

**Methods:** An anonymous questionnaire, developed in the UK in English and translated into local languages, was used in 2021-2022 to collect data on MMH and perceived coping ability from women aged ≥18 years with an infant born before or during lockdowns. Five MMH components (worry, sadness, loneliness, difficulty relaxing, annoyance) and coping were assessed on a 4-point Likert scale, then dichotomised. MMH and coping were compared across countries using Chi-square tests with post-hoc pairwise comparisons conducted via Bonferroni-adjusted z-tests. Predictors of MMH and coping were examined using multivariable logistic regression.

**Results:** A total of 7,650 women were analysed. Younger infant age, higher income, walking and exercise, and level of support were associated with better MMH and coping, whereas higher education was associated with better coping but poorer MMH. MMH and coping differed across countries (all p<0.001), which remained after adjusting for covariates: mothers in Asian countries reported better MMH, while those in the UK and Thailand reported better coping.

**Conclusions:** Postpartum MMH and coping during lockdown were shaped by both individual and contextual factors. Findings highlight cross-country differences and underscore the need to strengthen maternal support system during future disruptions to perinatal care.

**Key messages:**

- Consistent predictors of better mental health and coping included younger infant age, higher income, regular physical activity, and enough support-factors that are modifiable and relevant in future public health emergencies.
- Country of residence remained an independent predictor even after adjusting for individual and social factors, suggesting cultural norms, health service models, and policy responses play important roles.
- Although the COVID-19 pandemic is largely over, the findings offer important lessons for future crises. Strengthening support systems, promoting physical activity, and ensuring equitable caregiving within households can help protect postpartum mothers’ wellbeing during any major social disruption.

## Introduction

During the COVID-19 pandemic, government worldwide implemented various confinement or “lockdown” policies to curb transmission^1^. A *Lancet* article highlighted the urgent need to investigate the pandemic’s mental health impacts, particularly among vulnerable groups^2^. Women in the perinatal periods are one such group: even before COVID-19, up to 20% experienced perinatal psychiatric disorders^3^. Postpartum depression was of particular concern, with prevalence ranging from 6.9% to 12.9% in high-income countries and exceeded 20% in some low- or middle-income countries^4^.

Lockdowns may have adversely affected perinatal mental health^5^. The disruption of social support, especially after childbirth when families rely heavily on care and assistance, posed significant challenges^3,5^. Prevalence of depressive symptoms measured by the Edinburgh Postnatal Depression Scale (EPDS) at least doubled compared to pre-lockdown levels in several countries, reaching 28.6% in Italy, 23.6%. in Belgium, and 30% in China^6^. Women from ethnic minority backgrounds and socioeconomically deprived groups were disproportionately affected^3,7–9^.

Evidence on postpartum women’s mental health and coping during COVID-19 remains limited and largely drawn from single-country studies, restricting cross-national comparisons. Although several multinational studies have examined perinatal mental health using cross-sectional designs^10–12^, none has differentiated post-partum women, including those who were breastfeeding, from pregnant women.

Although the COVID-19 pandemic has ended, its impact on vulnerable groups, such as breastfeeding mothers and infants, remain important. Future public health emergencies will require culturally and socially tailored strategies informed by prior experience. This study therefore aimed to identify predictors of MMH and coping among new mothers during COVID-19 lockdowns, focusing on maternal characteristics, circumstances, and support factors, and to examine whether country of residence independently influenced these outcomes.

## Methods

### Study Setting and Design

Mothers aged 18 years or older living in the UK, China, Japan, Malaysia, Mexico, Argentina and Thailand, who had a young infant (≤12 months in the UK, Thailand, and Mexico; ≤24 months in Japan; ≤18 months in other countries) born before or during the lockdown/the State of Emergency in each country, were invited to complete a one_time, anonymous survey online or on paper. Data collection began on May 27, 2020 in the UK and continued until August 30, 2022. Ethical approval was obtained in all participating countries (eTable 1). Anonymous data from each country were combined by the research team at University College London (UCL).

### Questionnaire Design

A structured questionnaire was developed by a group of experts in Paediatrics, Child Nutrition and Dietetics, and Anthropology at UCL. It was first used in the UK COVID-19 New Mum Survey^9^ and subsequently translated into the local language of the collaborating countries with adaptations to reflect local context. The questionnaire collected information on maternal and infant background characteristics (including socio-economic status, living conditions, and support received), mother’s daily activities, mental health, and perceived coping ability during lockdowns (see supplementary questionnaire). For ethical reasons, no formal depression or anxiety assessments were conducted, as sensitive questions about suicidal ideation or self-harm could not be followed by appropriate support in an anonymous survey. Participants gave informed consent at the start of the survey and could choose to skip any questions they preferred not to answer.

### Data Reprocessing

Implausible values (for example, gestational age <21 or >43 weeks^13,14^) and invalid and indeterminate responses were reclassified as missing data. Maternal age, education, marital/household status, household income, and housing type were recoded to ensure comparability across countries (eTable 2). Based on principal component analysis (PCA) from previously published New Mum Study in the UK ^9^ and China^3^, we select five items to represent MMH:worry, sadness, loneliness, relaxation, and annoyance; and one item to represent coping: feeling able to cope with the situation.

### Outcome Recoding

Responses to MMH and coping questions, originally measured on a 4-point Likert scale, were recorded into binary outcomes by grouping “not at all” and “very little” versus “to some extent” and “to a high extent” (Table 1).

**Table 1.** Recording rules of outcome variables.

| Questions measuring outcomes | Recoding based on 4-point Likert scale |  |
| --- | --- | --- |
|  | Better<br>MMH/coping=1<br>(e.g., low level of<br>worry, high level of<br>coping) | Worse MMH/coping=0<br>(e.g., high level of worry, low<br>level of coping) |
| Worry: I've been feeling worried | Not at all/Very little | To some extent/To a high extent |
| Sadness: I've been feeling down | Not at all/Very little | To some extent/To a high extent |
| Loneliness: I've been feeling<br>lonely | Not at all/Very little | To some extent/To a high extent |
| Relaxation: I've had trouble<br>relaxing | Not at all/Very little | To some extent/To a high extent |
| Annoyance: I've become easily | Not at all/Very little | To some extent/To a high extent |
| Coping: I feel able to cope with the<br>situation | To some extent/To a<br>high extent | Not at all/Very little |

### Statistical Analyses

Background characteristics was described by country using number (percentage) for categorical variables or mean ± standard deviation (SD) for continuous variables. The distribution of MMH and coping was described by country and compared using Chi-square tests, with post-hoc pairwise comparison performed using Bonferroni-adjusted z-test. Multivariable logistic regression models were used to examine the strength of association between MMH, coping, and their predictors, with separate models fitted for each outcome. Predictors were identified using a directed acyclic graph (DAGitty version 3.0, eFigure 1) and included country, frequency of daily activities, support received, maternal age and education, infant age, gestational age, household income, marital/household status, living conditions, primiparity, and whether birth occurred before or during the lockdown. Odds ratio were reported alongside 95% confidence intervals (CI). Data from Argentina were excluded from this analysis because data on household income and support were not collected. All analysis was conducted in SPSS version 27 (IBM., Armonk, NY, USA).

**Figure 1.**
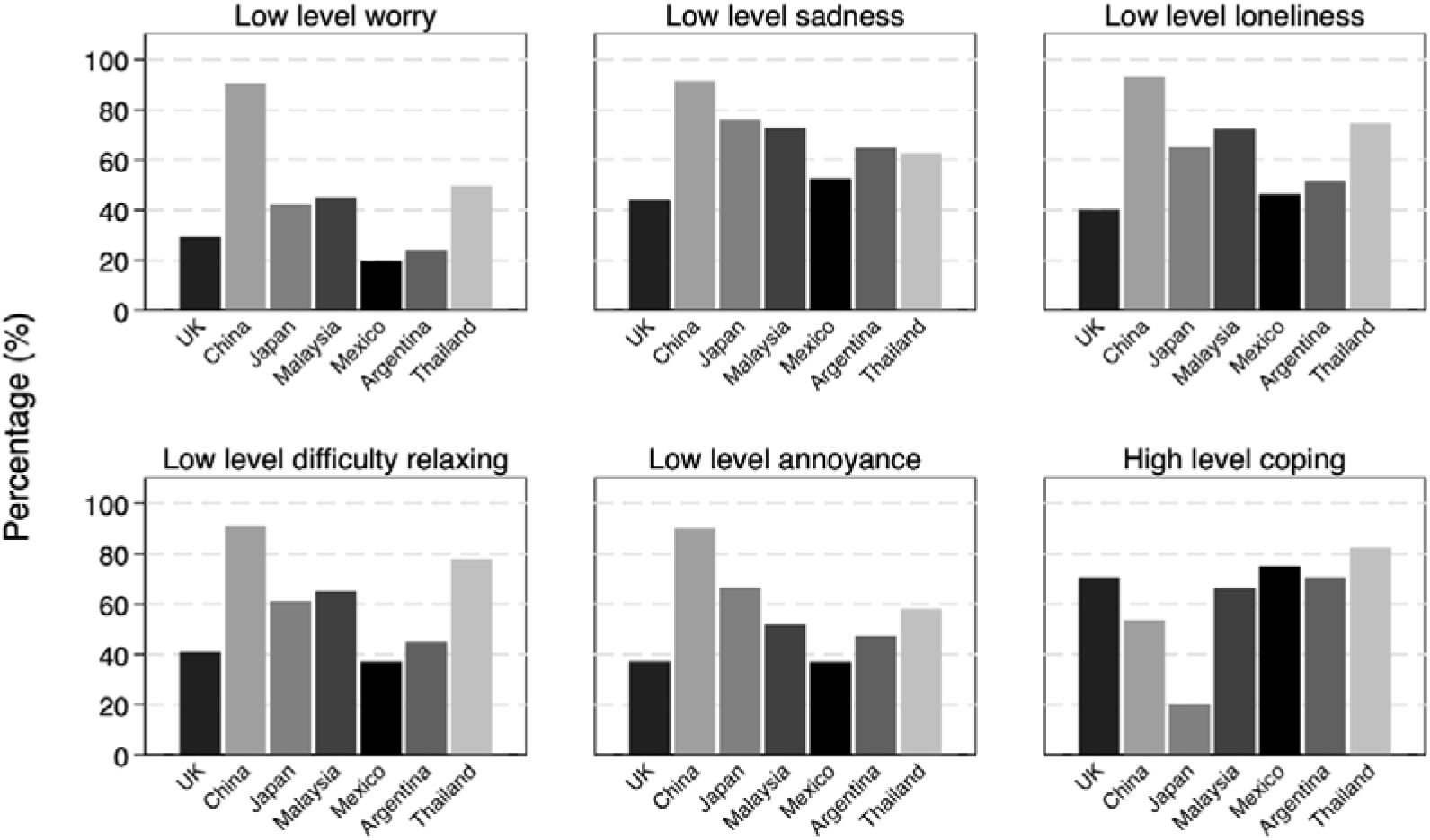
Percentage of participants reporting better MMH and coping by countries with results of post-hoc pairwise comparison. Notes: Chi-square test was conducted with post-hoc pairwise comparison of column proportions using Bonferroni-adjusted z-tests. P<0.05 was considered a statistically significant difference. All pairwise comparisons were significant apart from the following: 1) Low level of worry: UK-Argentina, Japan-Malaysia, Japan- Thailand, Malaysia-Thailand, Mexico-Argentina; 2) Low level of sadness: Malaysia-Argentina, Argentina-Thailand; 3) Low level of loneliness: UK-Mexico, Malaysia-Thailand, Mexico-Argentina; 4) Low level of difficulty relaxing: UK-Mexico, UK-Argentina, Mexico-Argentina; 5) Low level of annoyance: UK-Mexico, Japan-Malaysia, Japan- Thailand, Malaysia- Argentina, Malaysia-Thailand; 6) High level of coping: UK-Malaysia, UK-Mexico, UK-Argentina, Malaysia-Argentina, Mexico-Argentina.

## Results

### Background Characteristics

Of the 9,019 participants enrolled, 1,369 were excluded for not meeting the inclusion criteria (maternal age <18 years, infant age >18 months, birth after lockdown, or outside the study country during lockdown) or for missing key information, leaving 7650 participants from seven countries for inclusion in the analysis. Most participants were married (95.8%) and aged 26–35 years (69.9%). The majority reported having access to a suitable place to feed their infant (93.2%) and to green space (81.6%). Sufficient support was reported by 78.9% of mothers, whereas only 40.3% indicated higher levels of shared housework (eTable 3).

### MMH and Coping

More than half of participants reported experiencing low level of worry, sadness, loneliness, difficulty relaxing, annoyance and high level of feeling able to cope during lockdown (eTable 4). MMH and coping varied significantly across study countries (Figure 1, eTable 4, all p<0.001). Most country pairs showed significant differences in MMH and coping (p<0.05), except for a small number of pairs involving Mexico, Argentina, and Thailand. For instance, China showed a significant difference from all other countries in six outcomes: worry, sadness, loneliness, difficulty relaxing, annoyance, and perceived ability to cope (all p<0.05).

### Predictors of MMH

Results of the multivariable logistic regression of predictors of MMH during lockdown are shown in Figure 2 (A-C) and eTable 5. Several predictors showed consistent association across MMH outcomes. Women with higher income, enough support, residing in not overcrowded home were more likely to report low level of worry, sadness, loneliness, difficulty relaxing, and annoyance.

**Figure 2.**
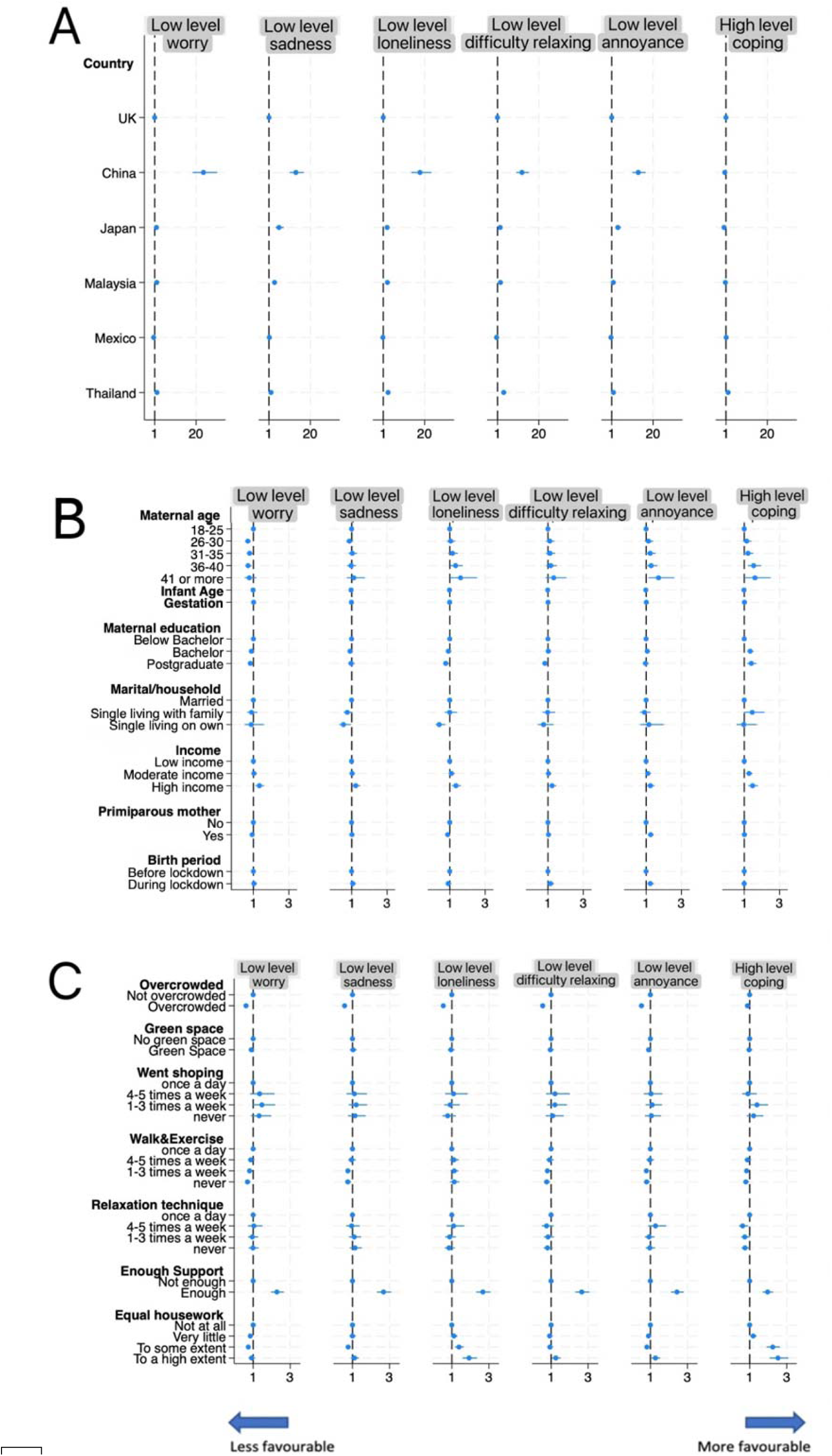
Predictors of maternal mental health and coping. Notes: Each plot presents adjusted odds ratios (ORs) with 95% confidence intervals (CIs) for predictors of MMH and coping during lockdown. Results are based on logistic regression models including all variables shown in panels A, B, and C, i.e., variables are mutually adjusted. Separate models were fitted for each outcome. Predictor selection was informed by a Directed Acyclic Graph (DAG) constructed using DAGitty (eFigure 1).

Compared with women on low income, women with higher income had 33% (OR=1.33, 95% CI: 1.11-1.58), 25% (1.35, 1.05-1.50) 36% (1.36, 1.13-1.63), 27% (1.27, 1.06-1.52), and 26% (1.26, 1.06-1.50) higher odds of reporting low level of worry, sadness, loneliness, difficulty relaxing, and annoyance, respectively. Women receiving sufficient support were more than twice as likely to report low level of worry, with OR ranging from 2.35 to 2.77 across MMH outcomes. Women who engaged in daily walking or exercise, compared with those exercising less than three times a week, were more likely to report low level of worry, sadness, difficulties relaxing, and annoyance.

Maternal age under 25, having a younger infant, and less shared housework were associated with low level of worry during the lockdown. Low level of sadness was associated with having a younger infant, while moderate shared housework was associated with higher level of sadness comparing with not shared housework. Married women reported a lower level of sadness than single mothers living on their own. Low level of loneliness was associated with maternal age above 35, having a younger infant, below bachelor’s degree, being married comparing with being single living on own, and more shared housework.

Predictors of low level of difficulty relaxing were below bachelor’s degree comparing with postgraduate, and high extent shared housework, while predictors of low level of annoyance included maternal age above 40, younger infant, primiparity, birth during lockdown, and high extent shared housework. Only moderate shared housework was associated with higher level of annoyance.

After adjusting for these predictors, differences in MMH remain significant across countries. Compared with women from the UK, levels of worry, sadness, loneliness, difficulty relaxing, and annoyance were lower among women in China, Japan, Thailand and Malaysia, but higher among women in Mexico (Figure 2A, eTable 5). Women in China showed 25.08 (19.99-31.46) times higher odds of reporting a low level of worry and 19.02 (14.92-24.24) times higher odds of reporting a low level of loneliness than women in the UK, In contrast, women in Mexico had 0.50 (0.38, 0.66), 0.57 (0.44, 0.74), and 0.63 (0.49, 0.83) times lower odds of reporting low levels of worry, difficulty relaxing, and annoyance, respectively, than women in the UK.

### Predictors of Coping

Figure 2 (A-C) and eTable 6 present predictors of feeling able to coping from a multivariable logistic regression model. Maternal age above 35, bachelor’s degree and above, being single living with family comparing with married, moderate income and above, not overcrowded residence, daily walking and exercise comparing with never exercise, enough support, and more shared housework to any extent were associated with better coping ability. For example, having a bachelor’s degree or postgraduate qualification was both associated with greater ability to cope (OR=1.31, 95% CI: 1.15, 1.50; OR=1.36, 95% CI: 1.14, 1.62). Single mothers living with family reported better ability to cope than married counterparts (OR=1.49, 95%CI:1.02, 2.17, p=0.039).

After adjusting for these predictors, country remained a significant predictor of coping (p<0.001). Compared with mothers in the UK, mothers in Japan (OR=0.12, 95% CI: 0.09, 0.16), China (OR=0.48, 95% CI: 0.40, 0.57), and Malaysia (OR=0.76, 95% CI: 0.62, 0.93) reported poorer ability to cope, whereas mothers in Thailand (OR=2.00, 95% CI: 1.52, 2.64) reported better coping, and no significant difference was found among mothers in Mexico (Figure 3A, eTable 6).

## Discussion

This large, cross-national study explored predictors of MMH and coping during the COVID-19 pandemic among over 7,600 postpartum mothers from seven countries. In general, reported MMH and coping varied significantly across countries, and younger infant, higher income, walking outside and exercise, and feeling of enough support consistently were associated with better MMH and coping. Higher education was linked to better coping but poorer MMH. Other potential predictors showed inconsistent associations across MMH and coping subdomains.

Younger infant age has been identified as a predictor of better mental health outcomes. A meta-analysis found that mothers of older infants reported significantly higher anxiety and depressive symptoms compared to those with younger infants, suggesting that cumulative caregiving demands may increase stress over time^15^. It is also possible that mothers with younger infants in our study may have had a greater focus on bonding and possibly received more social support and care in the early postpartum period during the pandemic.

Higher household income has been identified as a protective factor for MMH, both pre- and during the pandemic, as financial security likely reduces stress related to basic needs, housing, and access to childcare, thereby reducing psychological distress^16^. Additionally, higher income may increase access to private outdoor space, which is consistent with another finding in our study that walking outside and exercise more frequently predict better MMH and coping. Higher income may also be associated with better access with resources of relaxation methods such as yoga, online counselling, or mindfulness apps, which became more relevant during lockdowns^17,18^.

Regardless of the pandemic, higher levels of social support have consistently been associated with significantly lower rates of postpartum depression^19–21^. Our finding that maternal perceived social support and equitable sharing of household responsibilities during lockdown predicted better MMH and coping is consistent with these findings. Similarly, a cross_sectional study of over 2,300 postpartum mothers in the USA showed that higher perceived social support during lockdowns was associated with significantly lower rates of depressive and anxiety symptoms^22^. Looking forward, given that lockdowns limited in-person contact, alternative modes of support merit further investigation. A Chinese study during the pandemic demonstrated that participation in an internet support program improved maternal self_efficacy, reduced postpartum depression, and enhanced perceived social support in primiparous mothers ^23^. These findings suggest that innovation in support delivery could enhance maternal mental health and coping in times of social disruption, particularly online and community media–based approaches.

Notably, country of residence remained a strong independent predictor of MMH and coping in all models, even after adjusting for maternal, socioeconomic, and support-related factors. Mothers in China, Japan, Thailand, and Malaysia consistently reported more favourable mental health outcomes, yet mothers in the UK consistently reported better coping. These differences are likely multifactorial. In the case of China, it is possible that the prior experience of a nationwide lockdown during the 2003 SARS outbreak played a role^24^. This prior public health emergency may have prompted early development of remote psychosocial support systems and TV-based education during such special period^25^.

During the COVID-19 lockdown, Chinese mothers had access to various remote support formats, such as Dr. Cuiyutao Breastfeeding Q&A Platforms, community WeChat groups, livestreamed antenatal education, televised parenting programmes, community-based 24-hour helpline staffed by trained volunteers, joint parenting TV shows for couples, and late_night relaxation/music channels designed to reduce isolation, and foster coping. Although formal evaluations of these tools are limited, their availability may have buffered psychological stress. Comparatively, despite comparatively lower MMH, mothers from UK reported better coping among countries, reflecting their potential adaptation to help-seeking behaviours during lockdown. A qualitative study highlights how UK mothers framed lockdown as a “self-contained family unit,” with coping reinforced through online support groups and parenting communities^26^. Besides, as indicated by the UK part of this multi-country study, the predictors of better coping included support for their own health and more equitable division of household chores^27^.

However, beyond social response and services, cultural factors also warrant consideration. The top four countries with higher MMH (China, Japan, Thailand, and Malaysia) are East or Southeast Asian societies, where cultural norms often emphasise emotional restraint and mental health stigma remains prevalent^28,29^. Research has shown that individuals from East Asian cultures are less likely than those in Western societies to express emotions openly. They are also more inclined to conceal distress or to report psychological symptoms as physical complaints (known as somatization)^30,31^. As a result, mothers from those countries in our study may have been more likely to self-report more favourable outcomes even if their mental health was poor, potentially introducing bias.

This study is the first large cross-national analysis of maternal postpartum mental health and coping specifically during the COVID-19 lockdown period. The customised questionnaire captured a broad range of pandemic-related experiences, including infant feeding, perceived support, and mental health, which allows comprehensive and culturally relevant cross-country comparison.

However, several limitations should be acknowledged. First, the use of self-reported data introduces risks of recall bias, measurement error, and particularly social desirability bias. As discussed above, the willingness to report emotional distress may vary between cultures, potentially leading to underreporting in settings where mental health remains stigmatised. Second, the cross-sectional design limits causal interpretation, although potential confounders were adjusted for using a directed acyclic graph (DAG) approach. Third, variable recoding for harmonisation across countries may have reduced detail or introduced information bias. Fourth, sample representativeness was varied among participated countries. Finally, formal diagnostic tools for anxiety or depression were not used due to the anonymous survey format, which limits clinical interpretation.

## Conclusion

This study identified some consistent predictors of better maternal mental health and coping during lockdown across seven countries. However, cultural and national differences highlight the need for context-specific interpretation. Although the COVID-19 pandemic has passed, future public health emergencies are inevitable, and we must prioritise building scalable, inclusive support systems to protect maternal wellbeing in times of crisis. Future research should use longitudinal and mixed-method designs to explore how support systems and cultural norms influence maternal wellbeing, and to explore remote, cost-effective interventions for potential use of future public health emergencies.

## Conflict of Interest

Professor Mary Fewtrell receives an unrestricted donation for infant nutrition research from Philips (Amsterdam, NL). All other authors declare no conflicts of interest.

## Data Availability

The datasets generated and/or analyzed during the current study are not publicly available due to ethical and privacy restrictions but are available from the corresponding author on reasonable request.

## Acknowledgement

We would like to express our sincere gratitude to Dr. Mari Shinde for her valuable support in promoting the survey and facilitating participant recruitment in Japan.

## Declarations

### Funding

This project was funded by Professor Mary Fewtrell’s research funds in the Childhood Nutrition Research Group. Research support was provided by the NIHR Great Ormond Street Hospital Biomedical Research Centre.

### Ethics approval

Ethical approval was obtained in all participating countries. The study was approved by the UCL Research Ethics Committee, UK (0326/017); the Beijing Children’s Hospital Research Ethics Committee, China (2020-Z-102); the Human Research Ethics Committee of Kagawa Nutrition University, Japan (442); the Ethics Committee for Research Involving Human Subjects, Universiti Putra Malaysia, Malaysia (JKEUPM-2020-193); the Autonomous University of Yucatan Research Advisory Committee, Mexico (SISI/123/2021); and the Research Ethics Committee, Faculty of Medicine, Chiang Mai University, Thailand (COM-2563-07416). In Argentina, the study was conducted as part of a Master’s dissertation and did not require ethical approval according to local regulations.

### Consent to participate

Informed consent was obtained from all individual participants included in the study prior to participation.

### Consent for publication

No individual data or image is published.

### Code availability

Analyses were performed via the graphical user interface, no custom code was produced.

### Authors’ contributions

JY and MF designed and planed the study. MF, SD, ER, AV, JW developed the survey questionnaire. JY, SD, ER, AV, MK, KO, and NHMS coordinated and managed data collection. CL conducted data analysis under ML’s supervision. CL, ML, JY, and MF together drafted the manuscript. All authors revised the manuscript and approved it for submission.

## Notes

### Competing Interest Statement

The authors have declared no competing interest.

