## Supplemental Table 1 for "Predictors of maternal mental health and coping during the COVID-19 pandemic: A multi-country cross-sectional study"

**eTable 1** Ethical Approval

| Country | Ethical committee | Approval number |
| --- | --- | --- |
| UK | the UCL Research Ethics committee | 0326/017 |
| China | the Beijing Children’s Hospital Research Ethics committee | 2020-Z-102 |
| Japan | The Human Research Ethics Committee of Kagawa Nutrition University research committee | 442 |
| Malaysia | the Ethics Committee for Research involving Human Subjects, Universiti Putra Malaysia | JKEUPM-2020-193 |
| Mexico | Autonomous University of Yucatan research advisory committee | SISI/123/2021 |
| Argentina | Conducted as Master’s dissertation, no requirement for ethical approval | / |
| Thailand | the Research Ethics Committee, Faculty of Medicine, Chiang Mai University, Thailand | COM-2563-07416 |

**eTable 2** Recoding rules of all background characteristics

| Variables | UK | China | Japan | Malaysia | Mexico | Argentina | Thailand | Recoding |
| --- | --- | --- | --- | --- | --- | --- | --- | --- |
| Maternal age | Age in years | Age in years | Age in years | Date of birth | Age in years | Between 18 and 25 years | Age in years | Between 18 and 25 years |
|  |  |  |  |  |  | Between 26 and 30 years |  | Between 26 and 30 years |
|  |  |  |  |  |  | Between 30 and 35 years |  | Between 31 and 35 years |
|  |  |  |  |  |  | Between 35 and 40 years |  | Between 36 and 40 years |
|  |  |  |  |  |  | 40 years or more |  | 41 years or more |
| Maternal education | A-levels/High school diploma | Part-time undergraduate, associate degree or below | High school graduation | Secondary | Primary school | Tertiary level | Below lower secondary school | Below bachelor |
|  | 5 or more A-C grade GCSEs |  | Graduated from vocational school/junior college | Diploma/certificate | Secondary school | Secondary level | Lower secondary school |  |
|  | Less than 5 GCSEs A-C grade |  |  |  | High school |  | Upper secondary school |  |
|  |  |  |  |  |  |  | Diploma |  |
|  | Bachelor's degree | Full-time university degree | Graduate from university | Bachelor | University | Academic | Bachelor’s degree | Bachelor degree |
|  | Master's degree | Master’s degree | Completed master's course at graduate school | Masters | Postgraduate | Postgraduate (Specialization, Master's, Doctorate) | Master’s degree | Postgraduate and above |
|  | Doctoral or professional degree | PhD / Doctoral degree | Graduated from doctoral program | Doctorate/ PhD |  |  | Doctorate |  |
| Marital/household status | Married/Civil Partnership/Cohabitation | Married-living with husband | Married/same-sex marriage/cohabiting | Married | Married | Married/Concubinage: they all live together | Married/Concubinage | Married/Civil Partnership/Cohabitation-live with husband/partner |
|  |  | Married-living with husband and parents |  |  |  |  |  |  |
|  |  | Married-living with husband and parents-in-law |  |  |  |  |  |  |
|  |  | Married-living on own |  |  |  |  |  |  |
|  | Single mom- living with family |  | Single parent (living with parents or family) | Single mother - living with family members | Single living with her family | Single mom: lives with other family members | Single mom- living with family | Single mother-live with family |
|  | Single mom- living on own |  |  | Single mother - living alone | Single living just with her children | Single mother: lives alone with her child/children | Single mom- living on own | Single mother-live on own |
| Household income | Less than £20,000 | Less than 200,000 RMB/year | Less than 3 million yen | No income | Without income |  | Below 200,000 Baht | Low income |
|  | Less than £30,000 |  | 3 million yen or more - less than 5 million yen | Below RM 4000 | <= 1 minimum salaries |  | Below 500,000 Baht |  |
|  | Less than £45,000 |  |  |  | 1-1.9 minimum salaries |  | Below 800,000 Baht |  |
|  | Less than £75,000 | 200,000-300,000 RMB/year | 5 million yen or more - less than 7 million yen | RM4000 - RM7999 | 2-2.9 minimum salaries |  | Below 1,200,000 Baht | Moderate income |
|  |  | 300,000-400,000 RMB/year |  |  | 3-5 minimum salaries |  | Below 1,800,000 Baht |  |
|  | Less than £100,000 | 400,000-500,000 RMB/year | 7 million yen or more - less than 10 million yen | RM 8000 and above | >5 minimum salaries |  | Below 3,000,000 Baht | High income |
|  | Over £100,000 | Over 500,000 RMB/year | Over 10 million yen |  |  |  | Below 4,000,000 Baht |  |
|  |  |  |  |  |  |  | More than 4,000,000 Baht |  |
| Housing type | A mobile, temporary or other structure | High-rise apartment without a garden | Apartments and Condominiums | Apartment | Rent | Department | Flat or apartment | No attached independent outdoor space |
|  | A flat, maisonette or apartment | Slab-type apartment without a garden |  | Room for rent / Space for rent |  |  | Dormitory |  |
|  |  |  |  | Temporary room or other |  |  |  |  |
|  | A whole house or bungalow | High-rise apartment with a garden | Detached house | A house, terrace or bungalow | own | House | House | With attached independent outdoor space |
|  |  | Slab-type apartment with a garden |  |  |  |  |  |  |
|  |  | Villa with a garden |  |  |  |  |  |  |

**eTable 3** Background characteristics of mothers who completed the survey (mean ± SD/ N (%))

| Mean ± SD / N (%) | N (%) | Total  (N=7650) | UK  (N=1835) | China  (N=2092) | Japan  (N=425) | Malaysia  (N=1412) | Mexico  (N=787) | Argentina  (N=351) | Thailand  (N=748) |
| --- | --- | --- | --- | --- | --- | --- | --- | --- | --- |
| **Infant & maternal SES** |  |  |  |  |  |  |  |  |  |
| Maternal age (years) | 7650 (100) |  |  |  |  |  |  |  |  |
| 18-25 |  | 756 (9.9) | 148 (8.1) | 136 (6.5) | 15 (3.5) | 181 (12.8) | 171 (21.7) | 15 (4.3) | 90 (12.0) |
| 26-30 |  | 2474 (32.3) | 538 (29.3) | 769 (36.8) | 129 (30.4) | 542 (38.4) | 253 (32.1) | 43 (12.3) | 200 (26.7) |
| 31-35 |  | 2877 (37.6) | 734 (40.0) | 849 (40.6) | 182 (42.8) | 441 (31.2) | 249 (31.6) | 138 (39.3) | 284 (38.0) |
| 36-40 |  | 1328 (17.4) | 361 (19.7) | 298 (14.2) | 84 (19.8) | 212 (15.0) | 102 (13.0) | 121 (34.5) | 150 (20.1) |
| 41 or more |  | 215 (2.8) | 54 (2.9) | 40 (1.9) | 15 (3.5) | 36 (2.5) | 12 (1.5) | 34 (9.7) | 24 (3.2) |
| Infant age (months) | 7650 (100) | 7.0 ± 4.2 | 4.8 ± 3.1 | 8.2 ± 3.9 | 12.1 ± 4.6 | 7.0 ± 4.6 | 6.5 ± 3.5 | 7.9 ± 5.3 | 6.2 ± 2.9 |
| GA (weeks) | 7556 (98.8) | 38.7 ± 1.7 | 39.3 ± 1.8 | 38.8 ± 1.5 | 38.8 ± 1.5 | 38.3 ± 1.6 | 38.3 ± 1.6 | 38.4 ± 2.2 | 38.4 ± 1.5 |
| Infant sex | 7646 (99.9) |  |  |  |  |  |  |  |  |
| Male |  | 3892 (50.9) | 919 (50.1) | 1058 (50.6) | 207 (48.8) | 698 (49.4) | 408 (51.8) | 205 (58.4) | 397 (53.1) |
| Female |  | 3745 (49.0) | 913 (49.8) | 1031 (49.4) | 214 (50.5) | 714 (50.6) | 379 (48.2) | 143 (40.7) | 351 (46.9) |
| Prefer not to say |  | 9 (0.1) | 3 (0.2) | 0 (0) | 3 (0.7) | 0 (0) | 0 (0) | 3 (0.9) | 0 (0) |
| Maternal education | 7605 (99.4) |  |  |  |  |  |  |  |  |
| Below bachelor |  | 2688 (35.3) | 548 (30.1) | 874 (42.3) | 161 (37.9) | 585 (41.4) | 158 (20.3) | 149 (42.5) | 213 (28.5) |
| Bachelor’s degree |  | 3445 (45.3) | 733 (40.2) | 876 (42.4) | 238 (56.0) | 630 (44.6) | 455 (58.3) | 139 (39.6) | 374 (50.0) |
| Postgraduate |  | 1472 (19.4) | 542 (29.7) | 316 (15.3) | 26 (6.1) | 197 (14.0) | 167 (21.4) | 63 (17.9) | 161 (21.5) |
| Marital/household status | 7599 (99.3) |  |  |  |  |  |  |  |  |
| Married |  | 7283 (95.8) | 1747 (96.3) | 2056 (99.7) | 419 (98.6) | 1391 (98.5) | 727 (92.4) | 326 (92.9) | 617 (82.5) |
| Single living with family |  | 233 (3.1) | 21 (1.2) | 0 (0) | 2 (0.5) | 18 (1.3) | 54 (6.9) | 12 (3.4) | 126 (16.8) |
| Single living on own |  | 74 (1.0) | 46 (2.5) | 0 (0) | 4 (0.9) | 3 (0.2) | 6 (0.8) | 10 (2.8) | 5 (0.7) |
| others |  | 9 (0.1) | 0 (0) | 6 (0.3) | 0 (0) | 0 (0) | 0 (0) | 3 (0.9) | 0 (0) |
| Household income | 7266 (99.5) |  |  |  |  |  |  |  |  |
| Low income |  | 2740 (37.7) | 600 (32.9) | 701 (33.8) | 104 (24.5) | 646 (45.8) | 135 (17.2) | / | 554 (74.1) |
| Moderate income |  | 2242 (30.9) | 588 (32.3) | 781 (37.7) | 130 (30.7) | 455 (32.2) | 172 (21.9) | / | 116 (15.5) |
| High income |  | 1545 (21.3) | 557 (30.6) | 159 (7.7) | 172 (40.6) | 311 (22.0) | 285 (36.3) | / | 61 (8.2) |
| Not specify |  | 739 (10.2) | 78 (4.3) | 432 (20.8) | 18 (4.2) | 0 (0) | 194 (24.7) | / | 17 (2.3) |
| Primiparous mother | 7629 (99.7) | 4447 (58.3) | 999 (54.6) | 1329 (64.0) | 312 (73.4) | 563 (39.9) | 523 (66.5) | 242 (68.9) | 479 (64.0) |
| Gave birth BL | 7647 (100) | 4627 (60.5) | 1338 (73.0) | 1292 (61.8) | 282 (66.5) | 865 (61.3) | 140 (17.8) | 176 (50.1) | 534 (71.4) |
| **Living Conditions** |  |  |  |  |  |  |  |  |  |
| Overcrowded | 7600 (99.3) | 2202 (29.0) | 669 (36.6) | 771 (37.5) | 126 (29.8) | 309 (21.9) | 113 (14.4) | 90 (25.6) | 124 (16.6) |
| Suitable place to feed | 7262 (99.5) | 6770 (93.2) | 1818 (99.1) | 1823 (88.5) | 398 (94.3) | 1337 (94.7) | 661 (84.0) | / | 735 (98.3) |
| Access to green space | 7613 (99.5) | 6213 (81.6) | 1797 (98.5) | 1637 (79.2) | 401 (94.4) | 1086 (76.9) | 607 (77.2) | 331 (94.3) | 354 (47.3) |
| Housing type^a^ | 6789 (98.9) | 4781 (70.4) | 1534 (84.1) | 1353 (66.6) | 145 (34.3) | 951 (67.4) | / | 229 (65.2) | 569 (76.1) |
| Pet | 7603 (99.4) | 2674 (35.2) | 908 (49.7) | 309 (15.0) | 61 (14.4) | 412 (29.3) | 469 (59.7) | 221 (63.0) | 294 (39.3) |
| **Level of support** |  |  |  |  |  |  |  |  |  |
| Enough support | 7166 (98.2) |  |  |  |  |  |  |  |  |
| Enough |  | 5656 (78.9) | 1066 (58.3) | 1801 (91.4) | 291 (68.8) | 1103 (78.1) | 715 (91.3) | / | 680 (90.0) |
| Not enough |  | 1510 (21.1) | 763 (41.7) | 170 (8.6) | 132 (31.2) | 309 (21.9) | 68 (8.7) | / | 68 (9.1) |
| Equal housework | 7540 (98.6) |  |  |  |  |  |  |  |  |
| Not at all |  | 2328 (30.9) | 668 (36.7) | 599 (29.0) | 221 (52.1) | 245 (17.4) | 152 (21.2) | 105 (29.9) | 338 (45.2) |
| Very little |  | 2168 (28.8) | 480 (26.3) | 725 (35.1) | 118 (27.8) | 398 (28.2) | 245 (34.2) | 67 (19.1) | 135 (18.1) |
| To some extent |  | 1978 (26.2) | 409 (22.4) | 610 (29.5) | 58 (13.7) | 427 (30.2) | 173 (24.1) | 101 (28.8) | 200 (26.8) |
| To a high extent |  | 1066 (14.1) | 265 (14.5) | 133 (6.3) | 27 (6.4) | 342 (24.2) | 147 (20.5) | 78 (22.2) | 74 (9.9) |

Notes: N(%)=number of responding subjects (responding rate); SES: social economic status; GA: gestation age; BL: before lockdown, comparable group: DL: during lockdown; Overcrowded: “do you feel that your living space has become more overcrowded during lockdown?”; Suitable place to feed: “Is there currently a suitable place in your home to feed your baby?”; Access to green space: recombined and recoded question: “Do you have access to a public green space within walking distance? Including parks, garden, square…”; Housing type: recombined and recoded question: categorized as a resident with/ without attached independent outdoor space; Enough support: “Do you feel that you got or are getting enough support and help with your own health?” ; Equal housework: “I feel the house chores are more equally divided among household members”; “/”: variables that excluded certain country/countries from the analysis due to absence of data on question; ^a^ presenting the percentages of residences with attached independent outdoor spaces, comparable group: residences without independent outdoor space.

**eTable 4** Number (%) of mothers reporting better MMH and coping outcomes by country

|  | UK  (N=1835) | China  (N=2092) | Japan  (N=425) | Malaysia  (N=1412) | Mexico  (N=787) | Argentina  (N=351) | Thailand  (N=748) | Total  (N=7650) | P |
| --- | --- | --- | --- | --- | --- | --- | --- | --- | --- |
| Low level of worry | 535 (29.3%) | 1874 (90.5%) | 180 (42.5%) | 636 (45.0%) | 143 (19.9%) | 84 (23.9%) | 372 (49.9%) | 3824 (50.7%) | < .001 |
| Low level of sadness | 803 (44.0%) | 1892 (91.4%) | 323 (76.0%) | 1027 (72.7%) | 377 (52.6%) | 228 (65.0%) | 468 (62.7%) | 5118 (67.8%) | < .001 |
| Low level of loneliness | 736 (40.2%) | 1930 (93.3%) | 275 (65.2%) | 1022 (72.4%) | 333 (46.4%) | 181 (51.6%) | 557 (74.7%) | 5034 (66.7%) | < .001 |
| Low level of difficulty relaxing | 746 (41.0%) | 1885 (91.1%) | 257 (61.0%) | 921 (65.2%) | 267 (37.2%) | 158 (45.0%) | 579 (78.0%) | 4813 (63.9%) | < .001 |
| Low level of annoyance | 682 (37.3%) | 1872 (90.2%) | 283 (66.6%) | 732 (51.8%) | 266 (37.1%) | 166 (47.3%) | 432 (58.1%) | 4433 (58.7%) | < .001 |
| High level of coping | 1293 (70.7%) | 1110 (53.7%) | 86 (20.2%) | 936 (66.3%) | 540 (75.3%) | 248 (70.7%) | 612 (82.4%) | 4825 (63.9%) | < .001 |

Notes: Chi-square test was conducted to display country difference, and results of post-hoc pairwise comparisons were described somewhere else (Figure 2); /: variables that excluded certain country/countries from the analysis because they did not include the question.

**eTable 5** Logistic regression on predictors of better MMH

| Variables (REF) | Low level of worry | Low level of sadness | Low level of loneliness | Low level of difficulty relaxing | Low level of annoyance |
| --- | --- | --- | --- | --- | --- |
|  | OR  (95 % CI) | OR  (95 % CI) | OR  (95 % CI) | OR  (95 % CI) | OR  (95 % CI) |
| **Infant & maternal SES** |  |  |  |  |  |
| Country (ref=UK) |  |  |  |  |  |
| China | **25.08 (19.99, 31.46) ***** | **13.99 (11.14, 17.57) ***** | **19.02 (14.92, 24.24) ***** | **12.46 (9.99, 15.53) ***** | **14.76 (11.88, 18.33) ***** |
| Japan | **1.87 (1.40, 2.50) ***** | **5.63 (4.10, 7.74) ***** | **2.98 (2.21, 4.03) ***** | **2.30 (1.72, 3.08) ***** | **3.92 (2.92, 5.27) ***** |
| Malaysia | **2.02 (1.65, 2.48) ***** | **3.57 (2.90, 4.40) ***** | **3.08 (2.49, 3.79) ***** | **2.42 (1.98, 2.97) ***** | **1.81 (1.48, 2.21) ***** |
| Mexico | **0.50 (0.38, 0.66) ***** | 1.11 (0.85, 1.44) | 0.87 (0.67, 1.13) | **0.57 (0.44, 0.74) ***** | **0.63 (0.49, 0.83) ***** |
| Thailand | **2.08 (1.64, 2.65) ***** | **1.90 (1.49, 2.43) ***** | **3.21 (2.47, 4.16) ***** | **3.97 (3.05, 5.16) ***** | **1.81 (1.42, 2.30) ***** |
| Maternal age (ref=18-25) |  |  |  |  |  |
| 26-30 | **0.67 (0.54, 0.82) ***** | 0.88 (0.71, 1.08) | 1.07 (0.86, 1.32) | 1.07 (0.87, 1.32) | 1.10 (0.89, 1.34) |
| 31-35 | **0.76 (0.61, 0.94) *** | 1.01 (0.81, 1.26) | 1.17 (0.94, 1.46) | 1.06 (0.85, 1.31) | 1.22 (0.99, 1.50) |
| 36-40 | **0.68 (0.53, 0.87) **** | 0.94 (0.73, 1.20) | **1.36 (1.05, 1.75) *** | 1.12 (0.87, 1.43) | 1.25 (0.99, 1.59) |
| 41 or more | 0.79 (0.53, 1.18) | 1.11 (0.73, 1.68) | **1.63 (1.05, 2.52) *** | 1.32 (0.87, 2.02) | **1.68 (1.13, 2.51) *** |
| Infant age (months) | **0.98 (0.96, 0.997) *** | **0.96 (0.95, .98) ***** | **0.98 (0.96, 0.997) *** | 0.98 (0.97, 1.00) | **0.98 (0.96, 0.996) *** |
| Gestational age (weeks) | 1.01 (0.98, 1.05) | 0.99 (0.95, 1.02) | 0.99 (0.96, 1.03) | 1.01 (0.97, 1.04) | 1.02 (0.99, 1.06) |
| Maternal education (ref=BB) |  |  |  |  |  |
| Bachelor’s degree | 0.89 (0.77, 1.03) | 0.93 (0.80, 1.07) | 0.87 (0.75, 1.01) | 0.99 (0.85, 1.14) | 1.05 (0.91, 1.20) |
| Postgraduate and above | 0.87 (0.72, 1.05) | 1.01 (0.83, 1.22) | **0.78 (0.64, 0.94) **** | **0.81 (0.67, 0.98) *** | 0.97 (0.81, 1.16) |
| Marital/household (ref=Married) |  |  |  |  |  |
| Single living with family | 0.86 (0.63, 1.19) | 0.78 (0.57, 1.06) | 0.96 (0.69, 1.33) | 0.92 (0.65, 1.29) | 0.95 (0.70, 1.29) |
| Single living on own | 0.92 (0.52, 1.63) | **0.55 (0.31, 0.95) *** | **0.41 (0.22, 0.76) **** | 0.76 (0.43, 1.32) | 1.16 (0.68, 2.01) |
| Income (ref=Low income) |  |  |  |  |  |
| Moderate income | 1.02 (0.87, 1.19) | 1.03 (0.88, 1.20) | 1.09 (0.93, 1.28) | 1.05 (0.90, 1.22) | 1.10 (0.95, 1.27) |
| High income | **1.33 (1.11, 1.58) **** | **1.25 (1.05, 1.50) *** | **1.36 (1.13, 1.63) ***** | **1.27 (1.06, 1.52) **** | **1.26 (1.06, 1.50) **** |
| Primiparous mother | 0.97 (0.85, 1.09) | 1.01 (0.89, 1.15) | 0.90 (0.79, 1.02) | 1.02 (0.90, 1.16) | **1.29 (1.15, 1.46) ***** |
| Gave birth DL (ref=BL) | 1.03 (0.88, 1.19) | 1.05 (0.90, 1.22) | 0.87 (0.75, 1.02) | 1.14 (0.98, 1.32) | **1.22 (1.06, 1.42) **** |
| **Living conditions** |  |  |  |  |  |
| Overcrowded (ref=No) | **0.61 (0.54, 0.70) ***** | **0.57 (0.50, 0.64) ***** | **0.54 (0.47, 0.62) ***** | **0.56 (0.49, 0.64) ***** | **0.51 (0.45, 0.58) ***** |
| Green space (ref=No) | 0.90 (0.76, 1.05) | 0.99 (0.84, 1.16) | 0.94 (0.79, 1.12) | 0.98 (0.83, 1.16) | 0.93 (0.79, 1.09) |
| **Daily activities (ref=daily or more)** |  |  |  |  |  |
| Went shopping |  |  |  |  |  |
| 4-5 times/ week | 1.29 (0.81, 2.03) | 1.01 (0.64, 1.61) | 0.93 (0.56, 1.56) | 1.11 (0.68, 1.81) | 1.03 (0.66, 1.61) |
| 1-3 times/ week | 1.45 (0.99, 2.13) | 1.17 (0.79, 1.71) | 0.84 (0.54, 1.30) | 1.14 (0.75, 1.72) | 1.20 (0.82, 1.75) |
| Never | 1.33 (0.90, 1.96) | 1.11 (0.75, 1.64) | 0.74 (0.47, 1.15) | 1.08 (0.71, 1.64) | 1.15 (0.78, 1.69) |
| Walk & Exercise |  |  |  |  |  |
| 4-5 times/ week | 0.89 (0.72, 1.09) | 0.94 (0.77, 1.16) | 1.10 (0.89, 1.36) | 0.91 (0.74, 1.11) | 0.99 (0.81, 1.21) |
| 1-3 times/ week | **0.80 (0.67, 0.96) *** | **0.75 (0.63, 0.89) ***** | 1.09 (0.91, 1.31) | **0.79 (0.66, 0.94) **** | **0.78 (0.66, 0.93) **** |
| Never | **0.68 (0.55, 0.83) ***** | **0.74 (0.60, 0.91) **** | 1.06 (0.85, 1.31) | **0.75 (0.61, 0.92) **** | **0.79 (0.65, 0.97) *** |
| Relaxation technique |  |  |  |  |  |
| 4-5 times/ week | 1.02 (0.71, 1.48) | 0.98 (0.68, 1.42) | 1.08 (0.71, 1.63) | 0.76 (0.51, 1.13) | 1.27 (0.88, 1.84) |
| 1-3 times/ week | 0.99 (0.75, 1.30) | 1.14 (0.86, 1.51) | 0.86 (0.63, 1.18) | 0.84 (0.62, 1.14) | 0.91 (0.69, 1.20) |
| Never | 1.05 (0.81, 1.36) | 1.23 (0.94, 1.60) | 0.84 (0.63, 1.13) | 0.81 (0.61, 1.08) | 0.94 (0.73, 1.22) |
| **Level of support** |  |  |  |  |  |
| Enough support (ref=No) | **2.35 (2.03, 2.72) ***** | **2.77 (2.41, 3.18) ***** | **2.68 (2.33, 3.08) ***** | **2.67 (2.32, 3.06) ***** | **2.44 (2.12, 2.81) ***** |
| Equal housework (ref=Not at all) |  |  |  |  |  |
| Very little | **0.85 (0.73, 0.99) *** | 1.00 (0.86, 1.16) | 1.05 (0.90, 1.22) | 0.89 (0.76, 1.04) | 0.89 (0.76, 1.03) |
| To some extent | **0.72 (0.61, 0.84) ***** | **0.75 (0.64, 0.87) ***** | **1.34 (1.14, 1.57) ***** | 0.89 (0.76, 1.04) | **0.77 (0.66, 0.89) ***** |
| To a high extent | 0.86 (0.72, 1.04) | 1.08 (0.89, 1.30) | **1.87 (1.54, 2.27) ***** | **1.22 (1.01, 1.46) *** | **1.22 (1.02, 1.46) *** |

Notes: Significant values were shown in bold font; CI=Confidence Interval; REF: Reference group; SES: Social-economic status; BB: Below bachelor; DL: During lockdown; BL: Before lockdown; IOS: Independent outdoor spaces; Went shopping: “Went shopping at the grocery store or pharmacy”; Walk & Exercise: “Went outside for a walk or for exercise”; Relaxation technique: “Practiced a relaxation technique”; prefer not to say/ others/ not specify in sex, marital/household status/ household income were excluded from the model. *p<0.05; **p<0.01; ***p<0.001.

**eTable 6** Logistic regression on predictors of better coping

| Variables (REF) | Feeling able to cope |
| --- | --- |
|  | OR  (95 % CI) |
| **Infant & maternal SES** |  |
| Country (ref=UK) |  |
| China | **0.48 (0.40, 0.57) ***** |
| Japan | **0.12 (0.09, 0.16) ***** |
| Malaysia | **0.76 (0.62, 0.93) **** |
| Mexico | 1.13 (0.86, 1.47) |
| Thailand | **2.00 (1.52, 2.64) ***** |
| Maternal age (ref=18-25) |  |
| 26-30 | 1.12 (0.92, 1.36) |
| 31-35 | 1.21 (0.99, 1.49) |
| 36-40 | **1.44 (1.13, 1.82) **** |
| 41 or more | **1.67 (1.10, 2.54) *** |
| Infant age | 0.99 (0.86, 1.11) |
| GA | 1.01 (0.97, 1.04) |
| Maternal education (ref=BB) |  |
| Bachelor’s degree | **1.31 (1.15, 1.50) ***** |
| Postgraduate | **1.36 (1.14, 1.62) ***** |
| Marital/household (ref=Married) |  |
| Single living with family | **1.49 (1.02, 2.17) *** |
| Single living on own | 1.01 (0.58, 1.77) |
| Income (ref=Low income) |  |
| Moderate income | **1.27 (1.10, 1.46) ***** |
| High income | **1.49 (1.24, 1.78) ***** |
| Primiparous mother | 0.96 (0.86, 1.09) |
| Gave birth DL (ref=BL) | 0.98 (0.86, 1.11) |
| **Living conditions** |  |
| Overcrowded (ref=No) | **0.84 (0.75, 0.95) **** |
| Green space (ref=No) | 1.02 (0.88, 1.19) |
| **Daily activities (ref=daily or more)** |  |
| Went shopping |  |
| 4-5 times/ week | 0.87 (0.58, 1.29) |
| 1-3 times/ week | 1.36 (0.97, 1.89) |
| Never | 1.14 (0.81, 1.59) |
| Walk & Exercise |  |
| 4-5 times/ week | 0.89 (0.74, 1,07) |
| 1-3 times/ week | 0.79 (0.68, 0.92) |
| Never | **0.72 (0.60, 0.88) ***** |
| Relaxation technique |  |
| 4-5 times/ week | 0.71 (0.49, 1.03) |
| 1-3 times/ week | 0.82 (0.62, 1.10) |
| Never | 0.85 (0.64, 1.11) |
| **Level of support** |  |
| Enough support (ref=No) | **1.94 (1.69, 2.23) ***** |
| Equal housework (ref=Not at all) |  |
| Very little | **1.23 (1.07, 1.41) **** |
| To some extent | **2.24 (1.93, 2.60) ***** |
| To a high extent | **2.58 (2.13, 3.13) ***** |

Notes: Significant values were shown in bold font; CI=Confidence Interval; REF: Reference group; SES: Social-economic status; BB: Below bachelor; DL: During lockdown; BL: Before lockdown; Went shopping: “Went shopping at the grocery store or pharmacy”; Walk & Exercise: “Went outside for a walk or for exercise”; Relaxation technique: “Practiced a relaxation technique”; prefer not to say/ others/ not specify in sex, marital/household status/ household income were excluded from the model. *p<0.05; **p<0.01; ***p<0.001.

**eFigure 1** Conceptual framework of predictors of MMH and coping ability

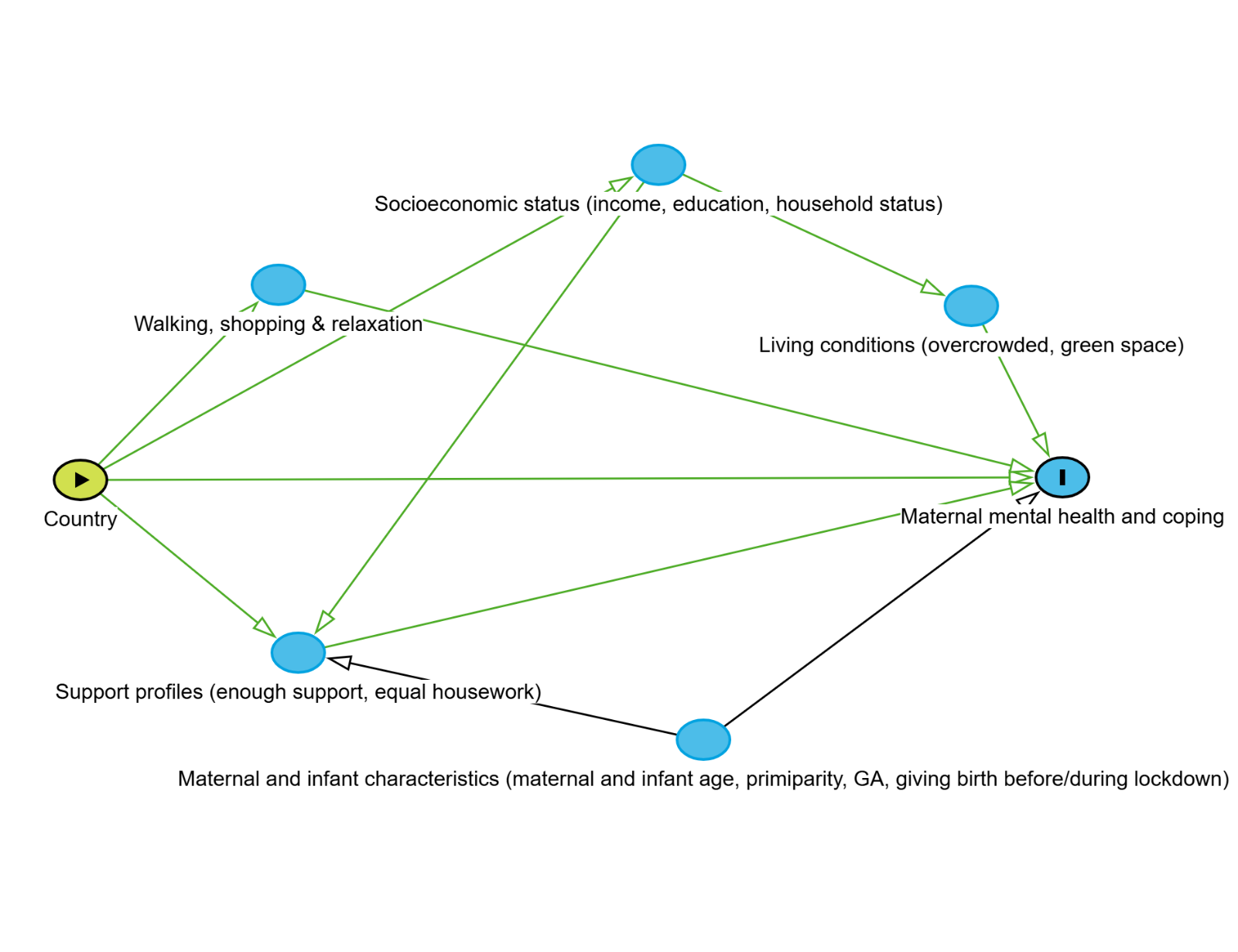
Notes: Country-exposure variable; Maternal mental health and coping-outcome variable; blue symbols without “I” presents covariates. The model was generated using DAGitty version 3.0 (online at <https://www.dagitty.net>)
